# Perspectives of professional experts in relation to the development of community-based exercise for young adults with schizophrenia – A qualitative study

**DOI:** 10.1101/2023.08.03.23293592

**Authors:** MF Andersen, K Roed, A Riis, BS Rafn, BH Ebdrup, J Midtgaard

**Affiliations:** Centre for Applied Research in Mental Health Care (CARMEN), Mental Health Centre Glostrup, University of Copenhagen, Copenhagen, Denmark; Department of Physiotherapy, University College of Northern Denmark, Aalborg, Denmark; Center for General Practice, Aalborg University, Aalborg, Denmark; Danish Cancer Society National Cancer Survivorship and Late Effects Research Center (CASTLE), Department of Oncology, Copenhagen University Hospital – Rigshospitalet, Copenhagen, Denmark; Center for Neuropsychiatric Schizophrenia Research (CNSR), Mental Health Centre Glostrup, University of Copenhagen, Glostrup, Denmark; Faculty of Health and Medical Science, Department of Clinical Medicine, University of Copenhagen, Copenhagen, Denmark

## Abstract

**Background:** Physical activity is a key modifiable factor in protecting physical and mental health in people with severe mental illness including schizophrenia. Therefore, early promotion of physical activity is recommended and programmes supporting long-term maintenance of physically active behaviour are warranted. This study aimed to explore the perspectives of professional experts in relation to the development of a sustainable community-based exercise programme tailored to young adults with schizophrenia and intended to promote change and adoption of physical activity.

**Methods:** We conducted nine semi-structured interviews with 11 clinical and professional experts. Qualitative content analysis, as described by Graneheim and Lundman, was applied to analyse data.

**Results:** We identified four categories: (1) living a physical active life with schizophrenia, (2) exercise as promotor of personal recovery, (3) prescribing safe and relevant exercise, and (4) instructors’ qualifications and formation.

**Conclusions:** When developing sustainable community-based exercise programmes tailored to young adults with schizophrenia, developers should ensure instructors’ qualifications and provide an exercise protocol. In addition, developers should consider providing an inclusive and recovery-oriented exercise environment.

**Key messages:** **What is already known on this topic**

- Physical activity is a key modifiable factor in protecting physical and mental health in people with severe mental illness, including schizophrenia.
- Sustainable community-based programmes to support adoption of physical activity for young adults with schizophrenia are warranted.

**What this study adds**

- Developers of community-based exercise for young adults with schizophrenia must ensure a strategy for identifying and training exercise instructors and the provision of a protocol for delivering safe and clinically relevant exercise.
- Community-based exercise may have the potential to promote personal recovery and thus considerations regarding the balance between overcoming potential barriers towards participation while promoting a non-clinical exercise environment are provided.

**How this study might affect research, practice or policy**

- These findings could support and inform the development of community-based programmes promoting physical activity for people with schizophrenia and may be adaptable or inspirable to other psychiatric populations in other geographical settings.

## Background

Severe mental illness, including schizophrenia, is among the most burdensome and costly illnesses worldwide (1). The clinical symptoms usually manifest in early adult life (2), and a substantial proportion of patients with schizophrenia experience persistent physical, functional, and cognitive impairments (3). Psychotic symptoms are often effectively treated with antipsychotic medication, however, treatment is commonly accompanied by metabolic side-effects. Patients treated with antipsychotic medication have a four-fold higher prevalence of metabolic syndrome (4), and a two-to-three-fold higher risk of cardiovascular disease compared to the general population (4), which contributes to a premature mortality rate of 15–20 years (5). Lifestyle factors, such as poor diet, smoking (6), and physical inactivity (7) may contribute to the increased morbidity and mortality.

The Lancet Psychiatry Commission has pointed to physical activity as a key modifiable factor in protecting physical health in people with mental illness (8). Specifically, introducing exercise in the early stages of schizophrenia may be a sustainable solution in preventing or attenuating metabolic dysfunction associated with antipsychotic medication (8,9). Furthermore, exercise has been found efficacious in improving clinical symptoms, quality of life, global functioning, and cognitive deficits (10–12), and early improvements in these outcomes may reduce the likelihood of enduring symptoms and functional disability (13). Moreover, patients in the early stages of schizophrenia are more physically active than patients with long-term schizophrenia (14), and thus potentially easier to engage in exercise. However, long-term maintenance of physical active behaviour is challenging (15).

Replicable and scalable methods of delivering physical activity to patients with schizophrenia in the early stages of their disease in a format that is accessible, engaging and effective for a large number are warranted (16). Indeed, the World Health Organization’s *Mental Health Action Plan 2013–2030* calls for the provision of mental health services integrated in communities for service users and families (*WHO, 2021*). Accordingly, community-based group exercise may hold promise to support sustainable physical activity (18). Still, the initiation of community-based exercise in people with serious mental illness is not a simple step from intention to participation, but a non-linear slow phased transition with various challenges and setbacks at every phase (19).

This study was conducted as a part of a project aiming to develop, evaluate and implement community-based exercise for young adults in antipsychotic treatment (ClinicalTrials.gov identifier: NCT05461885). Previously, we evaluated the impact and feasibility of a gym-based group exercise programme supervised by non-health professional exercise instructors for young adults in antipsychotic treatment (the COPUS trial) (20,21). The results indicated that gym-based group exercise has the potential to support and promote personal recovery, which can be defined as ‘*changing values, feelings, goals, abilities and roles in order to achieve a satisfactory, hopeful and productive way of life, with the possible limitation of the illnesś* (22). However, feasibility was challenged due to lack of incentives and infrastructure to ensure consecutively recruitment from outpatient clinics to gym-based exercise in communities. Thus it was concluded that refinements in the intervention and the delivery of the intervention were warranted before initiating a large-scale, definitive trial (20). Also, community-based exercise programmes may be considered a complex intervention due to the properties of the intervention itself, i.e., how the intervention produces change, and the interaction between the intervention and its context, i.e., how the context affects implementation and outcomes. Hence, early considerations and continually revisiting core elements, i.e., considering context, identifying key uncertainties, developing a programme theory, and refining the intervention, are recommended throughout the research process when evaluating complex interventions (23,24). The current study aimed to explore the perspectives of various professional experts in relation to the development of community-based exercise for young adults with schizophrenia intended to promote long-term maintenance of physical activity.

## Methods

### Design

We applied a descriptive qualitative design (25) using researcher triangulation and semi-structured interviews. This study follows the Consolidated Criteria for Reporting Qualitative Research (COREQ) checklist (26) (Supplemental Material 1).

### Sampling

We used a purposeful sampling strategy to support information richness (27). Specifically, we used snowball sampling to reach unique key informants with expert knowledge, experience, or interest concerning the study’s aims (28). Key informants were thus recruited among professional experts within exercise as health promotion and/or treatment of psychosis, e.g., psychiatry, physiology, exercise psychology. All key informants who were contacted agreed to participate. The final sample was defined by theoretical saturation, i.e., different expertise and perspectives represented, and by inductive thematic saturation during the analysis focusing on emerging new perspectives (29).

### Data collection

KR (female investigator, registered nurse, full-time PhD student) or MFA (male investigator, certified physiotherapist, full-time PhD student) carried out the semi-structured individual interviews, which lasted 45–60 minutes, using video calls on Microsoft Teams or physical face-to-face meetings. Only the interviewer and the key informants were present during interviews. Both KR and MFA have previous experience in conducting qualitative interviews and had no personal knowledge of the key informants beforehand. All key informants were provided with written or oral information regarding the study aim prior to the interviews. Hence, it was explained that community-based exercise was defined as group exercise delivered by non-health professional exercise instructors outside a hospital setting (e.g., commercial gyms or sporting clubs). The interview guide comprised a standard open-ended question:

> *According to you, as an expert, what should be considered when developing a community-based exercise programme for young adults with schizophrenia?*

In addition, each key informant was then asked specified questions thematically related to their area of expertise (Supplemental Material 2). The face-to-face interviews were audio-recorded, and the online video calls were video recorded. The interviewer (KR or MFA) who had conducted a specific interview wrote selective transcriptions, i.e., focusing exclusively on content relevant to the research question (30), which was then validated by the other researcher (KR or MFA) while focusing on and discussing the emergence of new perspectives to determine data saturation (29). To enhance trustworthiness, the key informants received the transcript within one week after the interview for member checking (31). Additional methodological considerations are available in Supplemental Material 2.

### Data analysis

The interviews were analysed using inductive qualitative content analysis, as described by Graneheim and Lundman (32). NVivo 12 (QRS International, Melbourne, Australia) and Microsoft Excel were used to assist in data management.

MFA and KR initially read and reread the transcripts separately before jointly discussing them to obtain a sense of the complete data material. Subsequently, MFA identified and extracted meaning units and labelled them with descriptive codes. Next, MFA compared, abstracted, and sorted codes into subcategories and categories, which were discussed with KR and JM (female psychologist, full-time researcher and principal investigator). Codes, subcategories, and categories were continuously compared with original condensed summaries in an iterative process. Figure 1 provides an example of the condensation-abstraction process.

**Figure 1.**
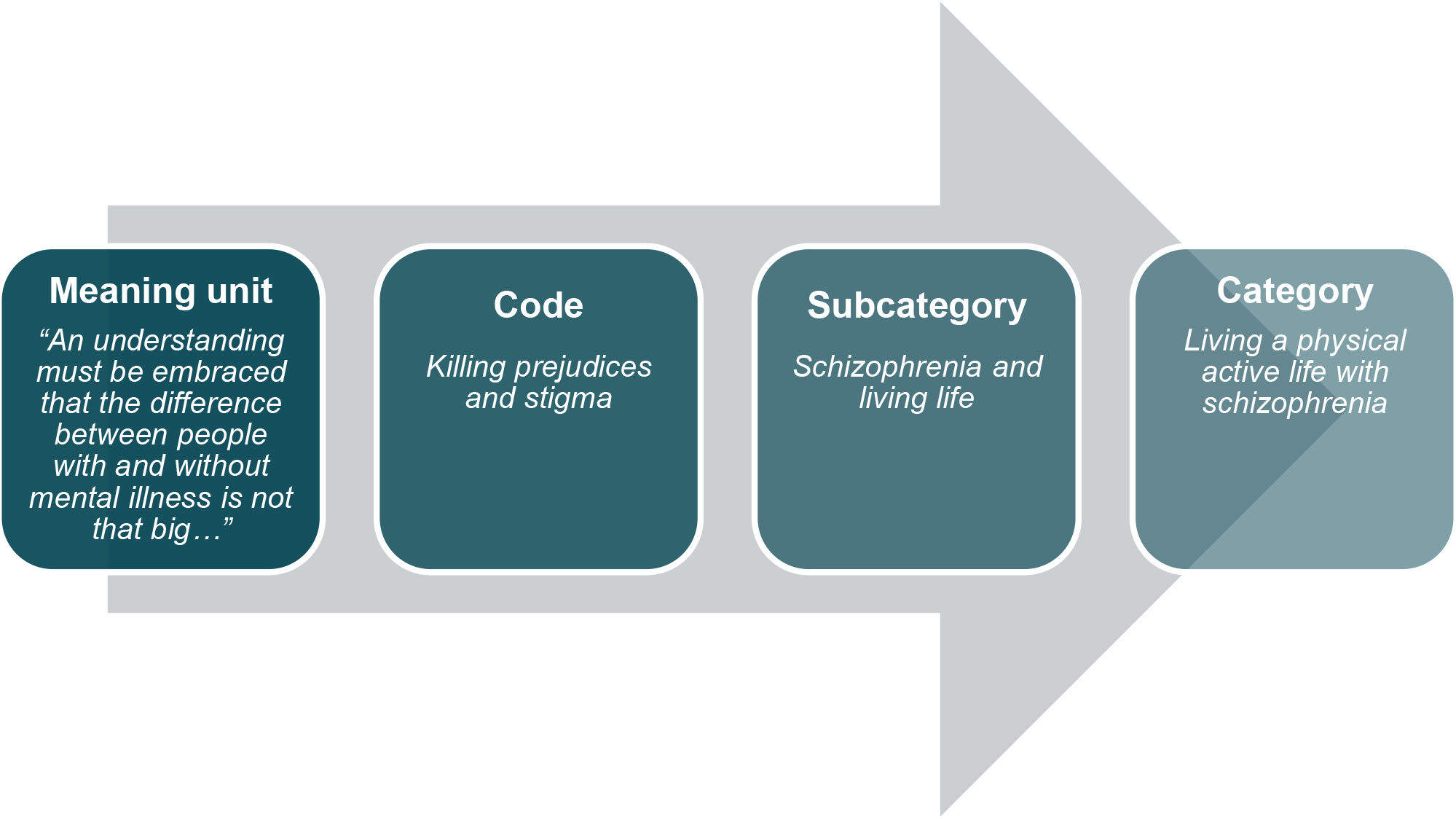
Example of the analytical process from meaning unit to category: *Living a physical active life with schizophrenia*.

### Ethics

In addition to receiving written information about the nature of the study prior to participation, all key informants provided oral informed consent. All key informants were guaranteed anonymity and confidentiality. Thus, quotes presented in the results section are not linked with the informant ID. The study was conducted in accordance with the Declaration of Helsinki. The Regional Ethics Committee of Northern Denmark has confirmed that no formal ethical approval was required (2023000206) for the current study.

## Results

### Characteristics of informants

Nine semi-structured interviews were conducted with 11 key informants (three key informants were interviewed simultaneously) representing different professional backgrounds i.e., psychiatry (n=1), psychology (n=4), human physiology (n=1), consultants with expertise in exercise for people with mental illness (n=2) and physiotherapy (n=2) and occupational therapy (n=1) within mental healthcare. All key informants had more than 10 years of experience working with people diagnosed with schizophrenia, exercise physiology and/or exercise psychology.

### Analysis of findings

Four main categories, 11 subcategories, and 34 codes were identified (Table 1). In the following presentation of the results young adults with schizophrenia will be addressed only as *participants* and non-health professional exercise instructors only as *instructors* as these were the terms used most frequently by the key informants.

**Table 1.**
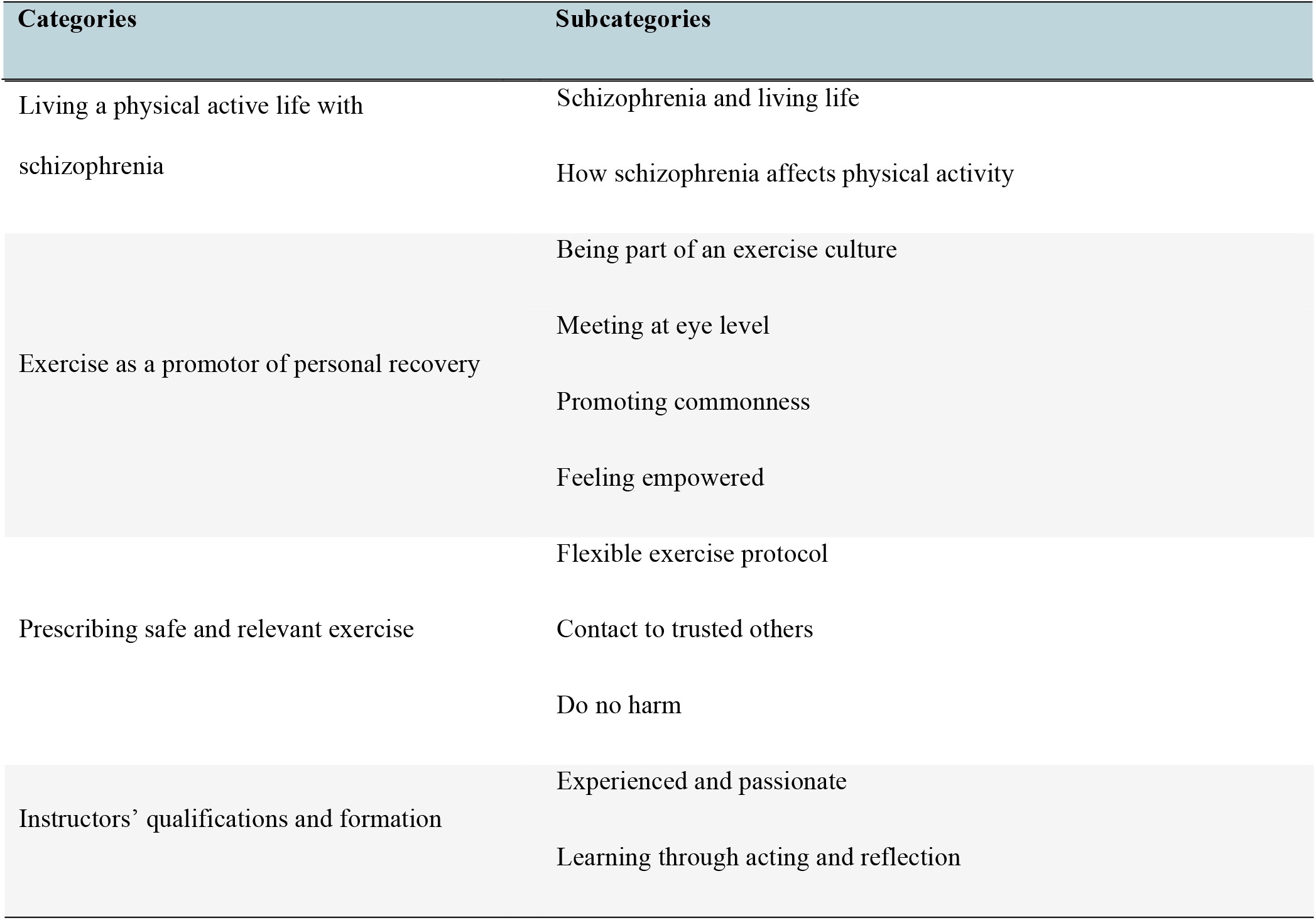
Results of the analysis.

### Category 1: Living a physical active life with schizophrenia

Schizophrenia is often associated with debilitating symptoms which have major impact on daily life and therefore also on the ability to engage in physical activities. The key informants emphasise that taking these potential barriers into consideration when developing a community-based exercise programme is vital along with ensuring that the exercise is supervised by instructors, who recognize these barriers.

#### Schizophrenia and living life

Suffering from schizophrenia may be perceived as a life crisis that may dampen hopes and dreams of living a “normal” life for the affected person. Community-based exercise may serve as a break from the disorder but may also constitute with yet another stressor:

> *Being struck by psychosis for a period in life can be exceedingly difficult… For some, physical exercise may be a ray of light that distracts them from the disease… But for others it may add to the list of things that have become unmanageable. (Expert in psychiatry)*

According to key informants with knowledge about mental illness, symptoms of schizophrenia may include a *psychophysical split*, implying that participants experience detachment from their own body. This may challenge participation in physical activity; however, exercise also has the potential to improve this phenomenon:

> *You can feel bodily disorganised and do not experience the body as one entity. There may also be the sensation that the body is levitating, and here physical exercising may be positive. (Expert in physiotherapy within mental healthcare)*

#### How schizophrenia affects physical activity

The key informants acknowledge that community-based exercise should be delivered by instructors, who have received basic education about schizophrenia, so they understand the participants’ potential reactions. However, the education must not facilitate to an overextended focus on the disease:

> *Must NOT dominate! Knowledge about this [psychotic disorders] should enable instructors to have realistic and informed expectations… But they should also know that people with mental illness are not significantly different from other people. (Expert in psychology and recovery)*

Especially, knowledge about psychotic and negative symptoms and how they may affect the participants’ ability to engage in physical activity with others is important, as the general population often misinterprets these symptoms.

> *Negative symptoms play an important role in the disease and are experienced as the most debilitating aspect associated with a schizophrenia diagnosis, in contrast to the general perception that it must be the psychotic symptoms… The intentions are present, but the initiative is absent. (Expert in psychiatry)*

Furthermore, severe weight gain is a typical side effect of antipsychotic medication which is often accompanied by considerable psychosocial distress. Several key informants caution, however, against weight loss becoming the most important goal as losing weight is difficult to achieve with physical activity alone. Instead, the experience of wellbeing and having a strong body should be promoted.

### Category 2: Exercise as a promotor of personal recovery

The key informants promoted the view that a community-based group exercise programme may have potential to facilitate participants’ personal recovery process. However, to achieve this potential, the programme should create an inclusive exercise environment.

#### Being part of an exercise culture

Of importance, the key informants stated that the participants should ideally achieve a feeling of being a part of something bigger than just an exercise programme; thus, participants should enter an exercise culture where being physically active is part of a regular everyday routine. To succeed, the setting must feel like a safe space where participants know the instructors and vice versa and atmosphere should reflect that all levels of participation are acceptable.

> *It must be a cool experience. If that succeeds… then the potential for physical activity has a chance to unfold. (Expert in psychology and physical activity for recovery)*

A safe atmosphere is created not only by the exercise instructor’s facilitation but more importantly within the group of participants. As a result, group interaction should be equally, if not more highly, prioritised compared to the content of the exercise. Nurturing social relationships between the participants may be challenged because of e.g., negative symptoms or anxiety; hence the instructor should take charge of facilitating a feeling of being a part of a group before, during, and after the exercise session. This may include having team exercise activities or encourage socialising after exercise sessions.

#### Meeting at eye level

The key informants expressed that it is important to have a caring communication without talking down to or being overprotective towards the participants. Participants may have experienced various disappointments and defeats regarding physical activity, potentially resulting in negative expectations and low self-confidence. Thus, self-stigma is considerable, and it must be acknowledged that even showing up may be a large accomplishment:

> *They [participants] will be tough enough on themselves if they only show up two out of three times. It’s important to say, “So nice to see you” rather than “Where were you last time?” Small things make the difference. (Expert in psychology and schizophrenia)*

#### Promoting commonness

One key informant underpins that the community-based exercise must not be a *sanctuary* for people with mental disorders disconnected from the rest of the world. Others state that being physically active where other young people are present and physically active might potentially be of great importance to the participants’ self-worth and in the (re)building of an exercise identity:

> *For a young patient to tell other young people that: “in my spare time I go to the gym three times a week” can be of great importance. (Expert in physiotherapy within mental healthcare)*

The participants should be met with the same demands and commitments as everyone else without violating their autonomy; it should be okay to withdraw for a moment if needed. There may be a general conception that people with schizophrenia are fragile and vulnerable, which may constitute a pitfall, and the exercise instructor may accidently take on a therapeutic role:

> *Instructors should listen but abstain from being therapeutic and giving advice [on mental health] and be aware of their task – which is exercise. (Expert in physiotherapy within mental healthcare)*

#### Feeling empowered

According to the key informants, a community-based exercise programme should seek to facilitate empowerment and draw attention to a positive sensation of being in control, being strong, having more energy, and sleeping better; in other words, of connecting exercise with a feeling of well-being. One key informant, however, emphasised the importance of recognising that exercise alone may not function as the main driver for all participants, but that they may find the context surrounding the exercise, such as the people and the place, more important to their well-being.

> *It may not be exercise as such making the difference, but the meaningfulness placed within the exercise. (Expert in exercise for people with mental illness)*

### Category 3: Prescribing safe and relevant exercise

Key informants agree that when developing a community-based exercise programme for young adults with schizophrenia, it is pivotal to ensure safety procedures in the case of physical and mental adverse events. Furthermore, the exercise protocol should have the flexibility to allow individual adjustments.

#### Flexible exercise protocol

Several key informants acknowledge that the exercise protocol should have a clear and recognisable structure. When the structure is known, varying the content and complexity of the exercise is easier for the instructors and more acceptable to the participants. It is important that the exercise content can be adjusted to fit the individual’s physical, mental, and social capacity.

> *It’s damn difficult because it’s not only about making the exercise easier or harder, but also about what the patient is like in the room and how they interact with the others. Instructors must acknowledge the complexity of an exercise situation. (Expert in physiotherapy within mental healthcare)*

The protocol must be flexible enough to accommodate individual goals of participants. However, even though individual goals may be mental or social, it is important that the instructors pursue clinically relevant exercise intensities given the considerably increased risk of metabolic diseases.

> *It’s important to pay attention to every single patient’s level of activity throughout the sessions, so each patient reaches a high level of activity over a longer period of time. (Expert in human physiology)*

#### Contact to trusted others

The key informants highlight that the support of both relatives and healthcare professionals is essential since they serve as external motivators and improve the ability of participants to attend the exercise sessions. Furthermore, inviting a friend or next of kin may ease the initial process of joining the group exercise. Because participants may express psychotic, suicidal, or aggressive behaviour during an exercise session, it is important that exercise instructors are provided safety procedures regarding what to do and who to contact. However, the key informants emphasise that such situations are very rare.

#### Do no harm

Some key informants express that elements of or being physically touched by others may be distressing for some participants. Furthermore, bodily discomfort such as delayed onset muscle soreness or an elevated heartbeat may be misinterpreted as dangerous and the normality of this needs to be addressed before, during, and after exercise sessions.

> *What can be expected after exercise, such as muscle soreness, and what can be expected during exercise, such as sweating, elevated heartbeat, and shortness of breath. Some of it may be confused with symptoms of anxiety. (Expert in physiotherapy in mental healthcare)*

### Category 4: Instructors’ qualification and formation

When developing a community-based exercise programme for young adults with schizophrenia, identifying the right exercise instructors is crucial. The key informants highlight that they should be fully engaged in their role as exercise instructors and receive formal training containing theory and the exchange of practical experience.

#### Experienced and passionate

According to the key informants, exercise instructors should be unpretentious and reliable to best support participants, who might be entering an unknown activity and culture. To do so, experience as an exercise instructor along with being passionate about this role is needed to focus on resonating with the participants. Furthermore, instructors should have relationship-building skills and be able to act as external motivation.

> *A motivational instructor is crucial for success. Without that, a good programme is worth nothing. (Expert in exercise psychology)*

#### Learning through acting and reflection

The key informants recommend that exercise instructors must receive formal training combining theoretical sessions as well as reflection on their own experiences to facilitate exercise for people with mental illness. Since instructors may have sparse experience and knowledge about mental illness, facilitating an ongoing exchange of experience is essential.

> *They [instructors] should receive more than just one day of training, actually a whole course, with the possibility to try something out, exchange experiences, and receive qualified feedback. (Expert in physiotherapy within mental healthcare)*

## Discussion

This qualitative study of the perspectives of clinical and professional experts offers a unique insight into core considerations when developing community-based exercise tailored to young adults with schizophrenia. It was widely acknowledged that mental health and social care initiatives in community-based settings are needed for people with severe mental illness to provide an opportunity to attain the highest level of health and participate in society free of stigmatisation and discrimination (33). Thus, key informants acknowledge that community-based exercise programmes have the potential to promote long-term maintenance of physical and mental health for young adults with schizophrenia, and thereby achieve the proposed potential of exercise (8,10–12)

Notably, our findings show a belief among experts in the potential of community-based exercise as an important contributor to personal recovery. They highlight that exercise could facilitate the experience of empowerment, i.e., of being strong and in control, which is an important factor in the recovery process according to one of the most accepted theoretical frameworks present to understand personal recovery, CHIME (Connectedness, Hope and optimism about the future, Identity, Meaning of life and Empowerment) (34). Furthermore, qualitative findings indicate that participating in sports contributes to experiencing a sense of achievement (35) and may represent an untapped resource in personal recovery as it can serve as an arena that promotes physically active behaviour while providing an opportunity to build life skills and social connectivity (36). Indeed, the key informants assert that group interaction should be prioritised at least as high as the content of the exercise. In addition, the community and the location should be a place that support the participants in transitioning beyond their role as patients to help them (re)build an exercise identity. Qualitative findings from our recent COPUS trial found that group exercise delivered in a conventional fitness centre by non-health professional exercise instructors supported a feeling of being a normal young adult (21).

The COPUS trial found that community-based exercise in a local gym was meaningful and safe for young adults with schizophrenia; however, it also reported that its feasibility was challenged due to limited recruitment and retention rates as the study setup did not provide the mental health staff with sufficient incitements and infrastructure to secure consecutive screening and promotion of the intervention (20). The key informants in this study highlight that support from trusted individuals, such as the primary healthcare provider, has an immense influence on the ability of people with schizophrenia to participate. This aligns with a qualitative review suggesting that emotional and practical support from either health professionals and/or relatives plays an important role (19). Moreover, while the exercise protocol should allow all levels of participation, clinically relevant exercise doses and intensities should be striven for to improve metabolic conditions. Current evidence suggests that higher doses of aerobic exercise are not only recommended to improve cardiovascular health but also clinical and functional outcomes (37).

Another important finding is the need for carefully recruiting dedicated exercise instructors as their personal engagement and ability to resonate with participants is crucial for success. This is in line with other developmental research on community-based physical activity programmes for people with mental illness and other clinical populations. Here, relationship-building skills, including empathy and the ability to generate team spirit are described as important instructor qualifications (19,38,39). The key informants in our study stressed the importance of instructors having formal education and knowledge on how symptomatology and the potential side effects of medical treatment may affect participation to allow them to have realistic and informed expectations as also supported by an international consensus statement (40). Especially negative symptoms, such as lack of energy, low self-esteem, depressive symptoms, and apprehensive attitude to socialising are the most frequently reported barriers toward exercise participation among people with schizophrenia (19,41). Furthermore, cognitive symptoms related to thinking about, planning and getting to an activity is also reported as a significant barrier (19). Knowledge about these symptoms may help instructors understand the importance of creating a welcoming and safe environment.

### Methodological considerations

This study has some strengths and limitations which should be taken into consideration. Despite our confidence in using purposeful sampling and snowball sampling aiming for variation in relation to the key informants’ areas of expertise, we cannot rule out an element of recruitment bias implying that key informants may have been positive about incorporating physical activity in community mental healthcare. Due to the geographical distance between key informants and the interviewers, most interviews were conducted using video calls. We found that this format was flexible, served the intended objective, and provided sufficient information power. For pragmatic reasons three informants who were colleagues were interviewed simultaneously. To account for potential conformity and authority bias the interviewer addressed specific questions to each key informant as representatives of their profession or area of expertise. Furthermore, each received the transcript for additional comments which were not shared with the other informants. Selective transcripts were used as empirical data, a process in which some relevant content may have been lost. Thus, to enhance trustworthiness, the transcripts were validated by a second researcher and sent to the experts for member checking.

### Clinical implications

Based on the perspectives of clinical and professional experts in the field, we propose the following recommendations encompassing structural and cultural factors in relation to promotion of long-term maintenance of physical activity in young adults with schizophrenia (Figure 2).

- Consider how schizophrenia may produce various barriers toward exercise participation and how paying special attention to them can be balanced with promoting a normal exercise environment.
- Consider how exercise may promote the personal and social recovery process, e.g., the CHIME framework, in the development of gym-based exercise programmes to support the sensation of empowerment, social connectivity, and exercise identity in participants.
- Ensure that safe and relevant exercise is prescribed, specifically by formulating a flexible exercise protocol and including other trusted individuals, e.g., relatives, friends, or mental health staff, to support participation and to agree on safety procedures.
- Ensure instructors’ qualifications and training, specifically by drawing up a strategy to identify and recruit experienced and passionate exercise instructors and by establishing an educational course and professional network that allows instructors to gain insight into schizophrenia and share reflection on practical experiences.

**Figure 2.**
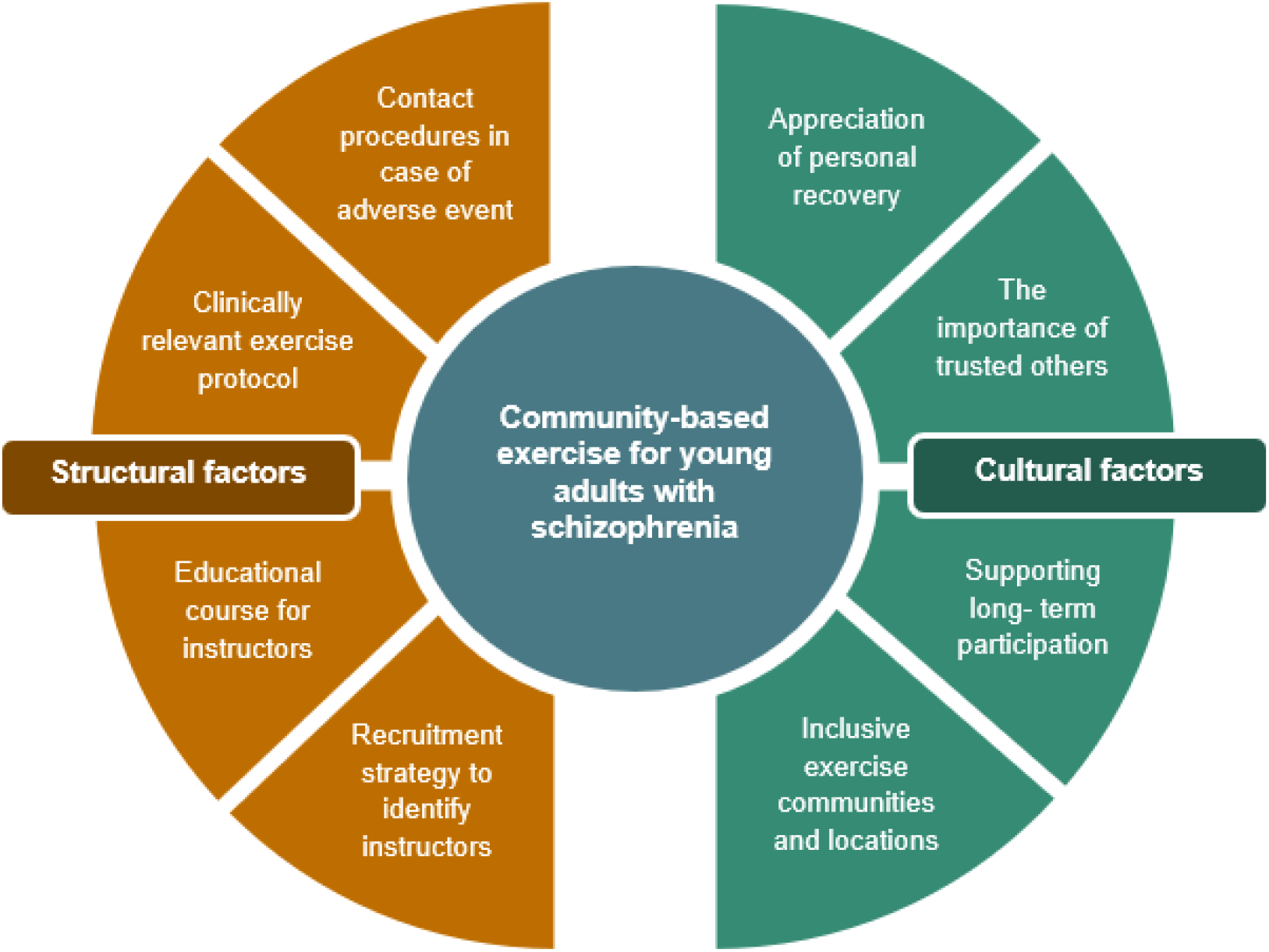
Structural and cultural factors of importance when developing community-based exercise targeting young adults with schizophrenia.

The current study was conducted in Denmark where the mental health care system and the community sector are closely interconnected to provide a comprehensive care to people with mental illness. However, we believe the recommendations from this study may be adaptable or inspirable to the development of programmes promoting physical activity in other psychiatric populations and in other geographical settings.

## Conclusion

This study provides key considerations when developing sustainable community-based exercise programmes tailored to young adults with schizophrenia intended to promote long-term maintenance of physical activity. Developers should focus on structural factors by ensuring instructors’ qualifications and providing a flexible exercise protocol, and cultural factors such as facilitating an inclusive and recovery-oriented exercise environment. These findings may be transferable to the development of programmes promoting physical activity in other psychiatric populations.

## Data Availability

All data produced in the present study are available upon reasonable request to the authors

## Acknowledgment

The authors are thankful to the key informants who participated in this study, for giving their time and providing detailed accounts of their expertise to make this work possible.

## Contributors

All authors participated in conceptualizing the study. MFA, KR and JM planned the study design and methodology. MFA and KR was responsible for the data collection and MFA was responsible for the initial data analysis. MFA, KR and JM contributed to the data analysis process. MFA generated the first draft, and all authors critically revisited the draft for important intellectual content. Lastly, the final version was sent to all authors for approval. All authors read and approved the final manuscript.

## Competing interests

BHE is part of the Advisory Board of Eli Lilly Denmark A/S, Janssen-Cilag, Lundbeck Pharma A/S, and Takeda Pharmaceutical Company Ltd; and has received lecture fees from Bristol-Myers Squibb, Boehringer Ingelheim, Otsuka Pharma Scandinavia AB, Eli Lilly Company, and Lundbeck Pharma A/S.

MFAN, KR, AR, BSR and JM declare no competing interests.

## Ethical approval

The Regional Ethics Committee of Northern Denmark has confirmed that no formal ethical approval was required (2023000206) for the current study. All key informants provided informed oral consent prior to participation in the study.

## Funding Source

The study was funded by TrygFonden (Grant No.: 151603)

## Notes

### Author Declarations

The Regional Ethics Committee of Northern Denmark has confirmed that no formal ethical approval was required (2023000206) for the current study.

