## Supplemental file 1 for "Perspectives of professional experts in relation to the development of community-based exercise for young adults with schizophrenia – A qualitative study"

**Supplemental Material 1** Consolidated criteria for reporting qualitative studies (COREQ): 32-item checklist

Please note that the COREQ checklist originally operates with reporting items according to which page number they can be found on. The page numbers in this supplementary file refer to the original manuscript. An added row to the checklist provides information as to which section and heading of the article specific items are reported in.

Developed from:

Tong A, Sainsbury P, Craig J. Consolidated criteria for reporting qualitative research (COREQ): a 32-item checklist for interviews and focus groups. *Int J Qual Heal Care*. 2007;19(6):349-357. doi:10.1093/INTQHC/MZM042

| **No. Item** | **Guide questions/description** | **Reported on Page #** | **Section and heading** |
|---|---|---|---|
| **Domain 1: Research team and reflexivity** | | | |
| *Personal Characteristics* | | | |
| 1. Inter viewer/facilitator | Which author/s conducted the inter view or focus group? | 6 | Methods under 'Data collection' |
| 2. Credentials | What were the researcher's credentials? E.g. PhD, MD | 6 | Methods under 'Data collection' |
| 3. Occupation | What was their occupation at the time of the study? | 6 | Methods under 'Data collection' |
| 4. Gender | Was the researcher male or female? | 6 | Methods under 'Data collection' |
| 5. Experience and training | What experience or training did the researcher have? | 6 | Methods under 'Data collection' |
| *Relationship with participants* | | | |
| 6. Relationship established | Was a relationship established prior to study commencement? | 6 | Methods under 'Data collection' |
| 7. Participant knowledge of the interviewer | What did the participants know about the researcher? e.g. personal goals, reasons for doing the research | 6 | Methods under 'Data collection' |
| 8. Interviewer characteristics | What characteristics were reported about the inter viewer/facilitator? e.g. Bias, assumptions, reasons and | 16 | Discussion under 'Methodological considerations' |

| | | | |
|---|---|---|---|
| | interests in the research topic | | |
| **Domain 2: study design** | | | |
| *Theoretical framework* | | | |
| 9. Methodological orientation and Theory | What methodological orientation was stated to underpin the study? e.g. grounded theory, discourse analysis, ethnography, phenomenology, content analysis | 7 | Methods under ‘Data analysis’ |
| *Participant selection* | | | |
| 10. Sampling | How were participants selected? e.g. purposive, convenience, consecutive, snowball | 6 | Methods under ‘Sampling’ |
| 11. Method of approach | How were participants approached? e.g. face-to-face, telephone, mail, email | 6 | Methods under ‘Sampling’ |
| 12. Sample size | How many participants were in the study? | 8 | Results under ‘Characteristics of informants’ |
| 13. Non-participation | How many people refused to participate or dropped out? Reasons? | 6 | Methods under ‘sampling’ |
| *Setting* | | | |
| 14. Setting of data collection | Where was the data collected? e.g. home, clinic, workplace | 6 | Methods under’ Data collection’ |
| 15. Presence of non-participants | Was anyone else present besides the participants and researchers? | 6 | Methods under ‘Data collection” |
| 16. Description of sample | What are the important characteristics of the sample? e.g. demographic data, date | 8,<br>Table 1 | Results under ‘Characteristics of informants’ |
| *Data collection* | | | |
| 17. Interview guide | Were questions, prompts, guides provided by the authors? Was it pilot tested? | 6<br>Appendix 2 | Methods under ‘Data collection’ |
| 18. Repeat interviews | Were repeat interviews carried out? If yes, how many? | 7 | Methods under ‘Data collection’ |
| 19. Audio/visual recording | Did the research use audio or visual recording to collect the data? | 6 | Methods under ‘Data collection’ |
| 20. Field notes | Were field notes made during and/or after the interview or focus group? | 7 | Methods under ‘Data collection’ |
| 21. Duration | What was the duration of the interviews or focus group? | 6 | Methods under ‘Data collection’ |
| 22. Data saturation | Was data saturation discussed? | 6, 7 | Methods under ‘Sampling’ and ‘Data collection’ |
| 23. Transcripts returned | Were transcripts returned to participants for comment and/or correction? | 6, 7 | Methods under ‘Data collection’ |
| **Domain 3: analysis and findings** | | | |
| *Data analysis* | | | |
| 24. Number of data coders | How many data coders coded the data? | 7 | Methods under ‘Data analysis’ |
| 25. Description of the coding tree | Did authors provide a description of the coding tree? | 8<br>Table 2 | Results under ‘Analysis of findings’ |
| 26. Derivation of themes | Were themes identified in advance or derived from the data? | 7 | Methods under ‘Data analysis’ |
| 27. Software | What software, if applicable, was used to manage the data? | 7 | Methods under ‘Data analysis’ |

| | | | |
|---|---|---|---|
| 28. Participant checking | Did participants provide feedback on the findings? | 7 | Methods under 'data collection' |
| *Reporting* | | | |
| 29. Quotations presented | Were participant quotations presented to illustrate the themes/findings? Was each quotation identified? e.g. participant number | 8-14 | Results under 'Analysis of findings' |
| 30. Data and findings consistent | Was there consistency between the data presented and the findings? | 8-14 | Results under 'Analysis of findings' |
| 31. Clarity of major themes | Were major themes clearly presented in the findings? | 8-14 | Results under 'Analysis of findings' |
| 32. Clarity of minor themes | Is there a description of diverse cases or discussion of minor themes? | 8-14 | Results under 'Analysis of findings' |
