## Supplemental file 2 for "Perspectives of professional experts in relation to the development of community-based exercise for young adults with schizophrenia – A qualitative study"

**Supplemental Material 2** Interview guide and additional methodological considerations

Standard opening and open-ended question:

*According to you, as an expert, what should be considered when developing a community-based exercise programme for young adults with schizophrenia?*

Expert specific question:

A:

- *With your practical experience in supervising exercise for people with schizophrenia, is there something you think non-health professional exercise instructors should pay close attention to? Motivation? Exercise content and structure?*

B:

- *With your knowledge of and experience in working with people with mental illness, what knowledge regarding pathophysiology should be shared with exercise instructors? Are there any contraindications that would inhibit people with psychotic disorders from participating in gym-based exercise? How should physical and mental complaints in relation to the exercise be handled?*

C:

- *With your knowledge of health psychology in relation to physical activity how would you create a motivating exercise environment outside the mental health system for people with mental illness? How do we avoid barriers towards change in physical active behavior?*

D:

- *With your practical experience in supervising exercise to people in antipsychotic treatment, is there something you think non-health professional exercise instructors should pay close attention to? Symptoms? Metabolic health?*

E:

- *With your experience in exercise programmes for people with mental illness, how would you create an inclusive and non-stigmatizing exercise environment? What should be considered in regard to the participants' social and personal recovery process?*

F:

- *With your knowledge of rehabilitation and recovery, what should be considered when delivering gym-based exercise to people in antipsychotic treatment in order to promote and support social and personal recovery? Are there any pitfalls that will work against recovery?*

G:

- *With your knowledge of and experience with physical activity in recovery processes, what would you consider when promoting exercise to people in antipsychotic treatment? Are there unique possibilities in using exercise instead of other social settings?*

H:

- *With your knowledge of exercise physiology, what should be considered to obtain clinically relevant exercise intensity and/or doses? How should the exercise content be organized? How should physical and mental complaints be handled during exercise?*

I, J, K:

- *With your knowledge of and clinical experience in working within early intervention outpatient clinics for people with schizophrenia spectrum disorders, how could an exercise community complement outpatient treatment?*

Additional methodological considerations

*We chose online video interviews (Microsoft Teams) due to the geographical distance between experts and researchers, as well as due to the assumption that sharing professional expertise would not evoke strong emotions. Furthermore, data elicited via video calls have demonstrated substantial richness akin to that gained in a face-to-face interview (1).*

*No fieldnotes were made during or after the interview and interviews were not repeated.*

*No patients took part in planning, conduct or interpretation of the results of this study.*

1. *Keen S, Lomeli-Rodriguez M, Joffe H. From Challenge to Opportunity: Virtual Qualitative Research During COVID-19 and Beyond. Int J Qual Methods [Internet]. 2022 May 1 [cited 2023 Jan 12];21:1–11. Available from: /pmc/articles/PMC9167989/*
